# Viral load dynamics in transmissible symptomatic patients with COVID-19

**DOI:** 10.1101/2020.06.02.20120014

**Authors:** Hitoshi Kawasuji, Yusuke Takegoshi, Makito Kaneda, Akitoshi Ueno, Yuki Miyajima, Koyomi Kawago, Yasutaka Fukui, Yoshihiko Yoshida, Miyuki Kimura, Hiroshi Yamada, Ippei Sakamaki, Hideki Tani, Yoshitomo Morinaga, Yoshihiro Yamamoto

## Abstract

To investigate the relationship between viral load and secondary transmission in novel coronavirus disease 2019 (COVID-19), we reviewed epidemiological and clinical data obtained from immunocompetent laboratory-confirmed patients with COVID-19 at Toyama University Hospital. In total, 28 patients were included in the analysis. Median viral load at the initial sample collection was significantly higher in adults than in children and in symptomatic than in asymptomatic patients. Among symptomatic patients, non-linear regression models showed that the estimated viral load at onset was higher in the index (patients who transmitted the disease to at least one other patient) than in the non-index patients (patients who were not the cause of secondary transmission; median [95% confidence interval]: 6.6 [5.2–8.2] vs. 3.1 [1.5–4.8] log copies/μL, respectively). High nasopharyngeal viral loads around onset may contribute to secondary transmission of COVID-19.

**Article Summary Line:** High nasopharyngeal viral load around the onset may contributes to secondary transmission of COVID-19.

## Introduction

Coronavirus disease 2019 (COVID-19), which is caused by the novel severe acute respiratory syndrome coronavirus 2 (SARS-CoV-2), has become a global pandemic, and currently threatens human health and lifestyles. Thus, it is important to gain an accurate understanding of the risk of infection with SARS-CoV-2. Generally, the risk of microbial human-to-human transmission is dependent on the duration of the highly-infectious phase and the number of virions contained in air particulates such as droplets and aerosols. However, as it is a new disease, little is yet known about COVID-19 and the risk of infection in various situations.

The viral load of SARS-CoV-2 peaks around the time of symptom onset, followed by a gradual decrease to a low level after about 10 days (*1,2*). Regarding the period of high infectiousness, a recent study reported that exposure to an index case within 5 days of symptom onset confers a high risk of secondary transmission (*3*). The high transmissibility around symptom onset gradually decreases, consistent with the dynamical pattern of viral shedding (*4*).

In addition, viral load can be associated with infectiousness, especially in the acute phase of COVID-19. However, little information is available on the relationship between nasopharyngeal viral load and secondary transmission. Therefore, in this study, we reviewed patients with COVID-19, including family clusters, and conducted follow-up interviews to investigate the relationship between viral load and secondary infection.

## Materials and methods

Epidemiological and clinical data were obtained from immunocompetent laboratory-confirmed patients with COVID-19 who were admitted to and/or from whom viral loads were measured at Toyama University Hospital. Index patients and those with secondary transmission were estimated based on serial intervals in the family clusters, and epidemiological and clinical data, including the number of secondary patients in health care, household, or other social settings, were investigated through structured telephone interviews. The study was performed in conformity with the Helsinki Declaration, after approval by the Ethical Review Board of University of Toyama (approval number: R2019167). Written informed consent was obtained from all patients.

The patients were divided into two groups: those who subsequently transmitted the disease to at least one other patient (index patients), and those who did not (non-index patients).

For each patient, the following data were retrieved from medical charts and structured telephone interview sheets: demographics, clinical presentation, date of symptom onset, date of initial sample collection, need for supplemental oxygen (moderate) and/or mechanical ventilation (severe), and dates of the first negative quantitative reverse transcription polymerase chain reaction (RT-qPCR) test result and hospital discharge.

Nasal swab specimens were pretreated with 500 μL of Sputazyme (Kyokuto Pharmaceutical, Tokyo, Japan). After centrifugation at 20,000 × g for 30 min at 4°C, the supernatant was used for RNA extraction. A total of 60 μL of RNA solution was obtained from 140 μL of the supernatant using the QIAamp ViralRNA Mini Kit (QIAGEN, Hilden, Germany) or Nippongene Isospin RNA Virus (Nippongene, Tokyo, Japan) according to the manufacturer’s instructions. The viral loads of SARS-CoV-2 were quantified based on an N2-gene-specific primer/probe set by RT-qPCR according to the Japan National Institute of Infectious Diseases protocol (5). The quality of quantification was controlled by AcroMetrix Coronavirus 2019 (COVID-19) RNA Control (Thermo Fisher Scientific, Fremont, CA). The detection limit was approximately 0.4 copies/μL (2 copies/5 μL).

Continuous and categorical variables were presented as the median (interquartile range [IQR]) and n (%), respectively. We used the Mann–Whitney U test, *χ*^2^ test, or Fisher’s exact test to compare differences between the index and non-index patients where appropriate. Data were analyzed using JMP Pro version 14.2.0 software (SAS Institute Inc., Cary, NC, USA). The viral load time courses were assessed using nonlinear regression employing a standard one-phase decay model in Prism version 8.4.2 (GraphPad Software Inc., San Diego, CA, USA).

## Results

Among the 28 patients (median age, 45.5 years) with laboratory-confirmed COVID-19, 15 (53.6%) were male, 21 (75.0%) were adults (18 years or older), and 10 (35.7%) were asymptomatic (Table 1). Among their 105 close contacts, 14 paired index-secondary cases were found. Fourteen (50.0%) were index patients within 11 family clusters, 10 (35.7%) were secondary transmission patients without further spreading within seven family clusters, and the remaining four (14.3%) were sporadic cases. Of the 18 symptomatic patients, the numbers of mild, moderate, and severe cases were 12, 5, and 1, respectively.

**Table 1.**
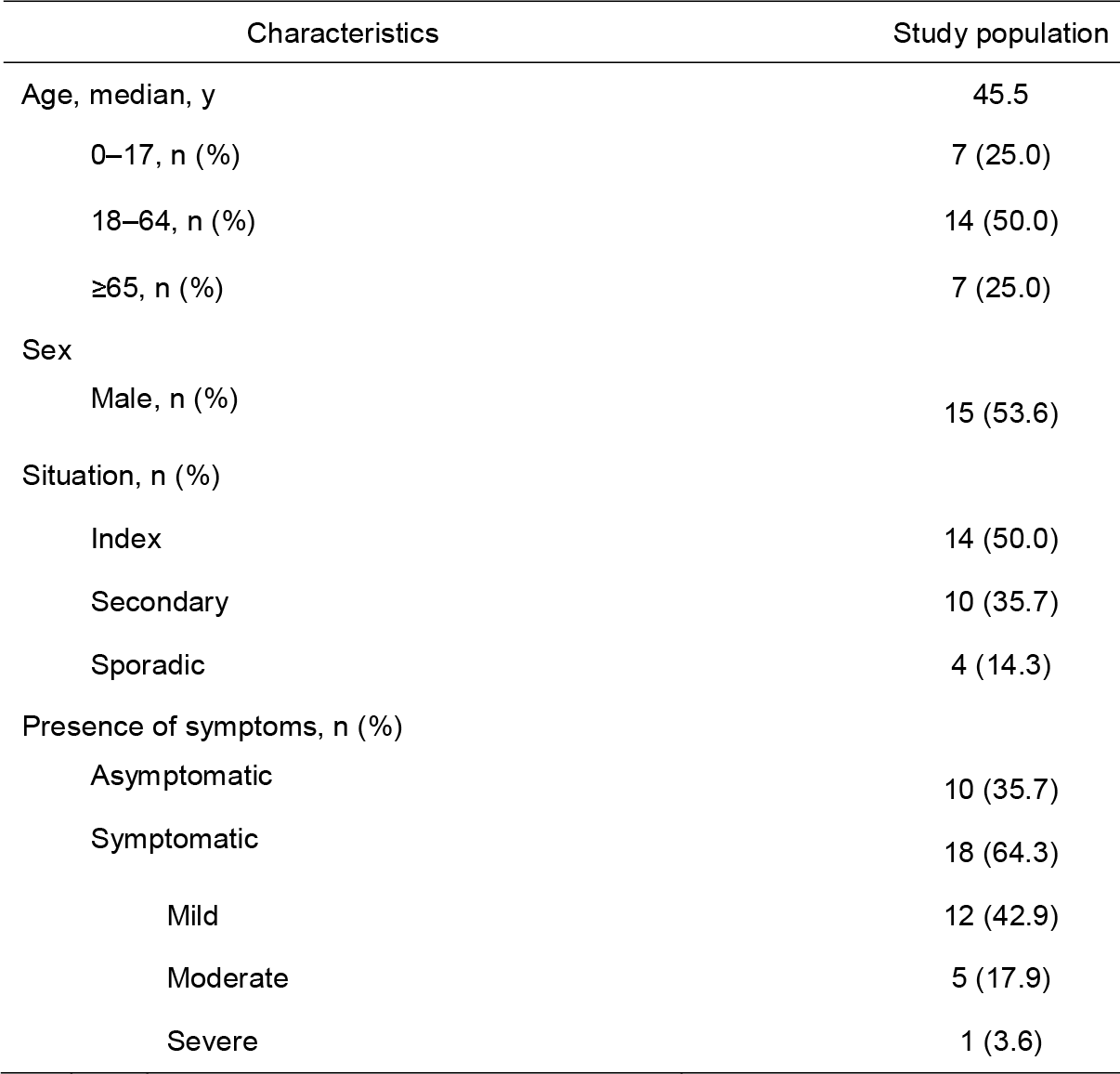
Basic characteristics of the patients.

A total of 89 nasopharyngeal swabs were collected from the 28 patients from 2 to 36 days after onset. The median (IQR) time at initial sample collection was 6 (2.8–9) days after symptom onset. The median viral load at the initial sample collection was significantly higher in adults than in children (p=0.02, 2.3 vs. 0.9 log copies/μL, respectively); however, viral loads during follow-up were not significantly different between the two groups (p=0.89). In addition, the median viral load at initial sample collection was significantly higher in symptomatic than in asymptomatic patients (p<0.01, 2.8 vs. 0.9 log copies/μL, respectively).

Next, the viral load in symptomatic patients was compared between the index and non-index patients (Table 2). Viral loads peaked soon after symptom onset, and then gradually decreased toward the detection limit (Figure 1A). The time to viral clearance from onset in the index patients was 21 (15–31) days (median [range]), and no significant difference was found between the index and non-index patients (p=0.09, 10 [9–26] days, median [range]).

**Figure 1.**
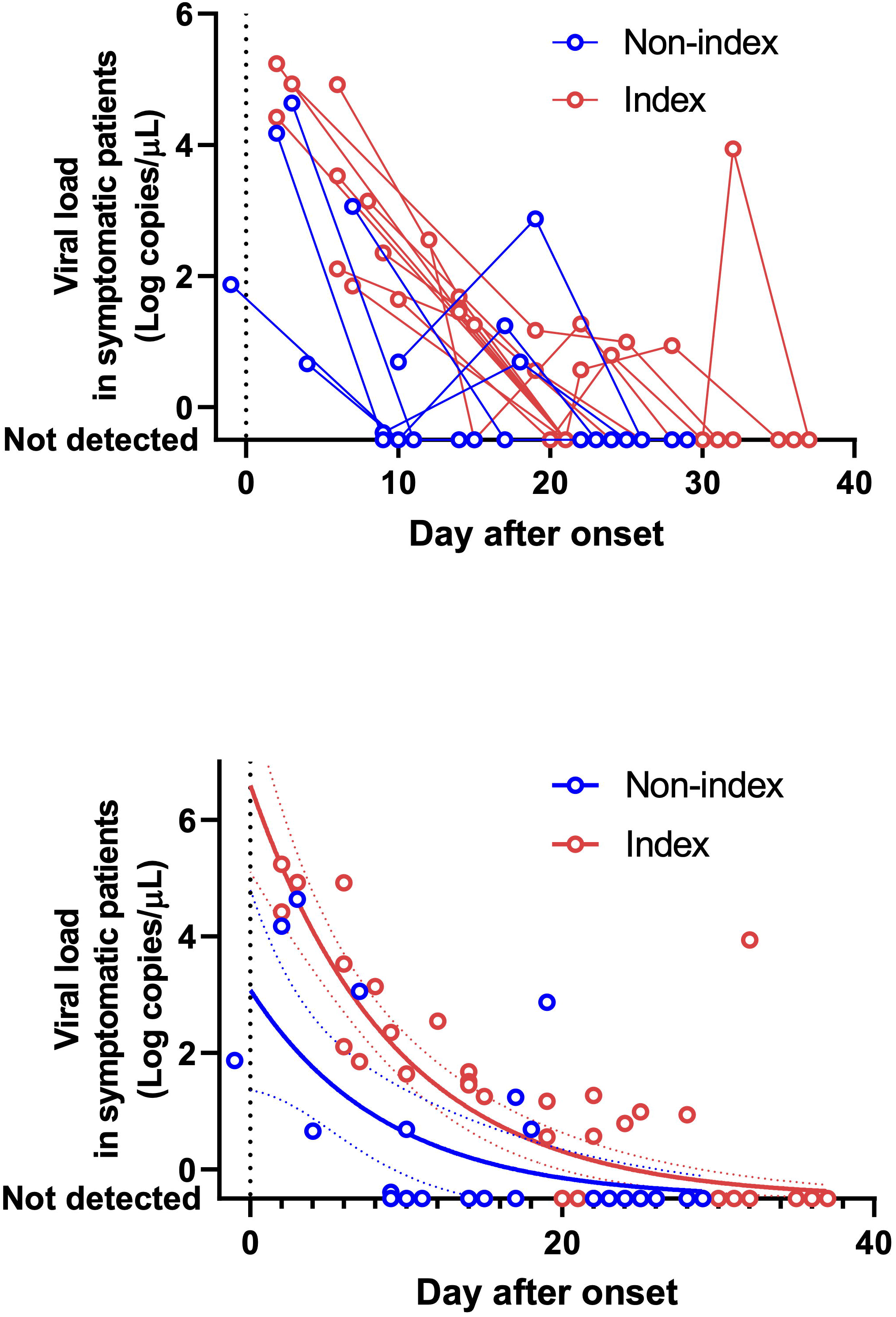
Trends in viral loads in the symptomatic patients. (A) Viral load time courses of index (red) and non-index patients (blue). (B) Nonlinear regression models of index (red) and non-index patients (blue). The models were calculated by using all the data of the index or non-index patients. When no virus was detected, the data were hypothetically plotted as –0.5 log copies/μL. Solid curves are best-fit models and dotted lines indicate the 95% confidence interval of each model.

**Table 2.** Summary of viral load in the index and non-index patients.

| Characteristics | Index patients,<br>n = 11 | Non-index patients,<br>n = 7 | P-value |
| --- | --- | --- | --- |
| Initial sampling |  |  |  |
| Sampling days after onset, median (range) | 6 (2 to 12) | 4 (–1 to 10) | 0.52 |
| Viral load (log copies/μL), median (range) | 3.1 (1.6 to 5.2) | 1.9 (–0.4 to 4.6) | 0.15 |
| Trend of viral load (log copies/μL), median (range) |  |  |  |
| –1 to 5 days after onset | 4.9 (4.4 to 5.2) | 3.0 (0.7 to 4.6) | 0.11 |
| 6 to 10 days after onset | 2.3 (1.6 to 4.9) | 0.7 (–0.4 to 3.1) | 0.17 |
| Days to viral clearance from onset, median (range) | 21 (15 to 31) | 11 (9 to 26) | 0.09 |

Among the symptomatic patients, the nasopharyngeal viral loads at the initial sample collection were not significantly different between the index and non-index patients (p=0.15, median [range]: 3.1 [1.6 to 5.2] vs. 1.9 [–0.4 to 4.6] log copies/μL, respectively). However, nonlinear regression models using all the data from the index or non-index patients showed that the viral load of the index patients at onset was higher than that of the non-index patients (median [95% confidence interval]: 6.6 [5.2 to 8.2] vs. 3.1 [1.5 to 4.8] log copies/μL, respectively), and this trend continued until 10 days after onset (Figure 1B).

## Discussion

In this study, we analyzed viral loads in nasopharyngeal swabs obtained from 18 symptomatic and 10 asymptomatic patients with laboratory-confirmed COVID-19 to assess the relationship between viral load and secondary transmission.

Although a recent study suggested that the viral load detected in asymptomatic patients was similar to that in symptomatic patients *(2*), in this study, the viral load at the time of initial sample collection was significantly higher in symptomatic than in asymptomatic patients. Furthermore, among the symptomatic patients, the viral loads in the index patients were significantly higher than those in the non-index patients at about 5 days after onset. Further studies are needed to confirm these results, as this study only involved a small number of patients. However, it is plausible that high nasopharyngeal viral loads contribute to secondary transmission of COVID-19. To our knowledge, no previous study has assessed the relationship between viral load and secondary transmission. Among the asymptomatic carriers, it was difficult to determine whether the viral load could impact transmission because no carrier had a high viral load.

After symptom onset, the viral load decreased monotonically. The median time to clearance of the virus was similar to that in a previous report, in which virus was detected for a median of 20 days (up to 37 days among survivors) after symptom onset (*6*). However, the median time did not differ between the index and non-index patients. These findings imply that viral clearance is independent of the initial virus load, but may be regulated by the host immune response. In most patients, the virus was detected for about 3 weeks; however, when the risk of transmission disappears remains unclear. A previous report demonstrated that infectiousness may decline at 8 days after symptom onset (*7*). Although the relationship between viral load and the infectiousness of COVID-19 remains unknown (*8*), the present study provides insight into the viral load threshold associated with infectivity.

This study has several limitations inherent to the small sample size and potential for confounding viral load and clinical conditions that cannot be excluded. The date of symptom onset and disappearance and information on disease transmission relied on self-reported information from the patients and their families, as well as information from public health centers, which could potentially lead to missing some cases of secondary transmission. In addition, the viral load dynamics were based on data from patients who received treatment, including combinations of antivirals and antibiotics, which could have modified the patterns of the viral load dynamics.

In conclusion, the results of this study suggest that a high nasopharyngeal viral load may contribute to the secondary transmission of COVID-19. In addition, the viral load may help explain why transmission is observed in some instances, but not in others, especially among household contacts. Although RT-qPCR does not distinguish between infectious virus and noninfectious nucleic acid, our findings may lead to the establishment of a viral load threshold to clarify COVID-19 disease transmission and infectivity.

## Data Availability

N/A

## Acknowledgment

We thank Ai Kakumoto for supporting PCR.

## Author Bio

H. Kawasuji is a clinical fellow of Toyama University Hospital. He is also a PhD-course student and has one publication about methicillin-resistant *Staphylococcus aureus* bacteremia as first author.

